# Foistar®(Camostat mesylate) associated with the significant decrease in CRP levels compared to Kaletra®(Lopinavir/Ritonavir) treatment in Korean mild COVID-19 pneumonic patients

**DOI:** 10.1101/2020.12.10.20240689

**Authors:** Jae-Phil Choi, Hyoung-Jun Kim, Jumi Han, SuJung Park, JinJoo Han

**Affiliations:** Seoul Medical Center, Seoul, South Korea; Daewoong Pharmaceutical, Seoul, South Korea

## Abstract

**Background:** There is limited information due to absence of virus titer and symptom related changes. Nonetheless, this is the first comparative study between the use of Foistar (Camostat mesilate) and Kaletra (lopinavir/ritonavir) on COVID-19 infection.

**Methods:** Patients with confirmed SARS-CoV-2 infection by positive polymerase chain reaction (PCR) testing that were admitted to Seoul Medical Center (Seoul, South Korea) where is the largest public medical center in South Korea between August 1 and September 20, 2020 were included

The data of the patients with pneumonia who received Foistar (Foistar group) during their hospitalization period were primarily collected, and the patients who received Kaletra (Kaletra group) during their hospitalization period were matched to have a similar age group to that of Foistar group so that three times the number of Foistar group patients were randomly selected into Kaletra group and their body temperature, CRP level, WBC count, and event of diarrhea were collected, accordingly.

**Results:** A total of 29 patients (7 Foistar group and 22 Kaletra group) was included. The median age was 69, and all had mild COVID-19 (WHO ordinal scale 3 or 4) on admission. 6 patients out of 7 patients (85.71%) from Foistar group who exhibited elevated CRP levels (CRP >0.4mg/dL) on admission have controlled their CRP levels to the normal range. In Kaletra group, 11 out of 18 patients (61.11%) have controlled their CRP levels to the normal range, and only 1 of 2 patients (50.00%) who had normal CRP level has maintained his or her normal CRP level. The difference in the white blood cell counts was not significant between two groups. None of the patients in the study had hyperkalemia.

**Conclusion:** This study has found a probable association of controlling inflammatory reactions and fever in COVID-19 patients with Foistar (camostat mesilate) use. In addition, there was no significant adverse drug event found from this study upon the Foistar use. These results may encourage the use of Foistar as a treatment option for the patients with mild to moderate COVID-19.

## Introduction

The new coronavirus infection (COVID-19), which began in December of 2019 in Wuhan, China, has spread worldwide. In Korea, the outbreak of patients has surged since February 24, 2020, and finally the World Health Organization (WHO) declared a pandemic on March 14, 2020. COVID-19 has a remarkable propagation power with the high basic reproduction number (R0) of 2-3, and the initial symptoms are non-specific resulting repeated clusters in the community. Currently, there is no effective vaccine or treatment that can suppress the epidemic, so it is very urgent to conduct clinical trials on drugs that can be expected to be effective among drugs developed previously.

SARS-CoV-2 is an RNA single-strand virus penetrating into the cells through the endosome by binding to the Spike protein, which is the structural protein, and the angiotensin-converting enzyme 2 (ACE2) receptor on the outer wall of the host inner cell. Type-2 Transmembrane protease (TMPRSS2) promotes priming of S-protein through proteolysis to induce binding to the cell membrane ACE2 receptor.1-3It synthesizes a polypeptide through viral RNA after entering a cell via an endosome. These polypeptides synthesize a RNA polymerase through the formation and activation of a replication-transcriptase complex, and replicate new RNA. The cloned RNA has a lifecycle that generates structural proteins and release them out of the cell again.

As described above, the virus penetrates into the cell through the binding of the spike protein and the ACE receptor, and in the meantime, TMPRSS2 plays a role in promoting the priming of the spike protein.^4^ Camostat mesilate can be used as an inhibitor of these TMPRSS2 to inhibit cell invasion of the virus.^5-10^

In lined with these assuring mechanisms of action of camostat against COVID-19, a total of 12 clinical trials have been registered in clinicaltrials.gov as of October 2020.^11-22^ The ongoing clinical trials are in the process to confirm the efficacy and safety of camostat in COVID-19 patients in the U.S., the UK, Denmark, Mexico, etc., along with South Korea. Accordinly, Daewoong Pharmaceutical’s Foistar Tab., which is composed of camostat mesilate, is expected to be effective in removing virus in COVID-19 patients and helping to relieve their symptoms. Thus, we conduct a retrospective cohort study to confirm the preliminary effectiveness of camostat use in patients with COVID-19 infection.

## Methods

### 1) Methods

We retrospectively reviewed medical records of COVID-19 pneumonic patients who confirmed the positive COVID-19 by the RT-PCR test result, and were admitted to Seoul Medical Center between August 1, 2020 and September 20, 2020. We included the patients who were administered with either Foistar (Foistar group) or Kaletra (Kaletra group) within 7 days from admission, and investigated its effectiveness and safety during the hospitalization period.

The body temperature, laboratory test results (i.e. CRP level, WBC counts, serum potassium level, etc.), and the number of patients who exhibited one or more events of diarrhea were compared between Foistar group and Kaletra group.

#### Body Temperature

The individual body temperature was recorded on a daily basis during the hospitalization period.

#### Laboratory test results

The records of CRP level, WBC count, serum potassium level of each patient were reviewed.

#### Event of Diarrhea

The number of patients who exhibited an event of diarrhea until discharge.

### 2) Statistical analysis

The collected data were analyzed via SAS®(Version 9.4 or above, SAS Institute Inc. NC. USA). The continuous variables (i.e. the number of patients, standard deviations, median, min, max) were presented as descriptive statistics, and the categorical variables were presented as a frequency, or a percentage.

The baseline in this study was defined as the day of admission. Any missing data in the day of discharge were replaced by the most current data before the discharge.

## Result

### 1) Patient Characteristics

A total of 29 patients were analyzed in this study. 7 patients (5 males, and 2 females) were included in Foistar group (camostat), and 22 patients (11 males, and 11 females) were included in Kaletra group (lopinavir/ritonavir). The mean age at the admission was 68.57 (±8.28) years in Foistar group, and 67.86 (±8.36) in Kaletra group. The mean of NEWS score upon the admission was 2.71 in Foistar group, whereas the 1.27 in Kaletra group.

Other characteristics such as WHO Ordinal scale, medical history, vital signs, respiratory rate, etc. are shown in table 1.

**Table 1.** Characteristics of the included patients.

|  |  | **Foistar Group**<br>(N=7) | **Kaletra Group**<br>(N=22) | **Total**<br>(N=29) |
| --- | --- | --- | --- | --- |
| **Sex, n(%)** | Female | 2 (28.57) | 11 (50.00) | 16 (55.17) |
|  | Male | 5 (71.43) | 11 (50.00) | 13 (44.83) |
| **Age\*(yrs)** |  | 68.57 (8.28) | 67.86 (8.36) | 68.03 (8.20) |
| **Hospitalization period\* (days)** |  | 15.29 (3.82) | 13.82 (1.50) | 14.17 (2.28) |
| **Duration of**<br>**Diagnosis~Hospitalization (days)** |  | 2.00 (1.15) | 2.05 (1.96) | 2.03 (1.78) |
| **WHO Ordinal scale Score, n(%)** | 3 | 5 (71.43) | 22 (100.00) | 27 (93.10) |
|  | 4 | 2 (28.57) | 0 (0.00) | 2 (6.90) |
|  | 5 | 0 (0.00) | 0 (0.00) | 0 (0.00) |
| **NEWS Score\*** |  | 2.71 (1.60) | 1.27 (0.98) | 1.62 (1.29) |
| **Medical History, n(%)** | Hypertension | 3 (42.86) | 6 (27.27) | 9 (31.03) |
|  | Diabetes | 1 (14.29) | 6 (27.27) | 7 (24.14) |
|  | Lung diseases | 0 (0.00) | 0 (0.00) | 0 (0.00) |
| **Systolic Blood Pressure\*(mmHg)** |  | 134.14 (20.94) | 142.36 (16.75) | 140.38 (17.81) |
| **Pulse\*(per min)** |  | 91.00 (8.77) | 94.55 (15.68) | 93.69 (14.26) |
| **Respiratory Rate\* (per min)** |  | 19.71 (1.38) | 18.68 (1.76) | 18.93 (1.71) |
| **Temperature\* (°C)** |  | 37.06 (0.85) | 36.95 (0.57) | 36.97 (0.64) |
| **Oxygen Saturation\* (%)** |  | 94.43 (1.62) | 96.23 (1.38) | 95.79 (1.61) |

SD = standard deviation, Min = minimum, Max = maximum, NC = not calculated.
Note: Denominator of percentage is the number of subjects in each group.
\* Mean(SD)

### 2) Body Temperature

The individual body temperature has been recorded on a daily basis and the number of patients who have displayed 38.0 °C or above were counted during their hospitalization period.

The trend of body temperature of an individual during the hospitalization period is presented below. (Figure 1)

**Figure 1.**
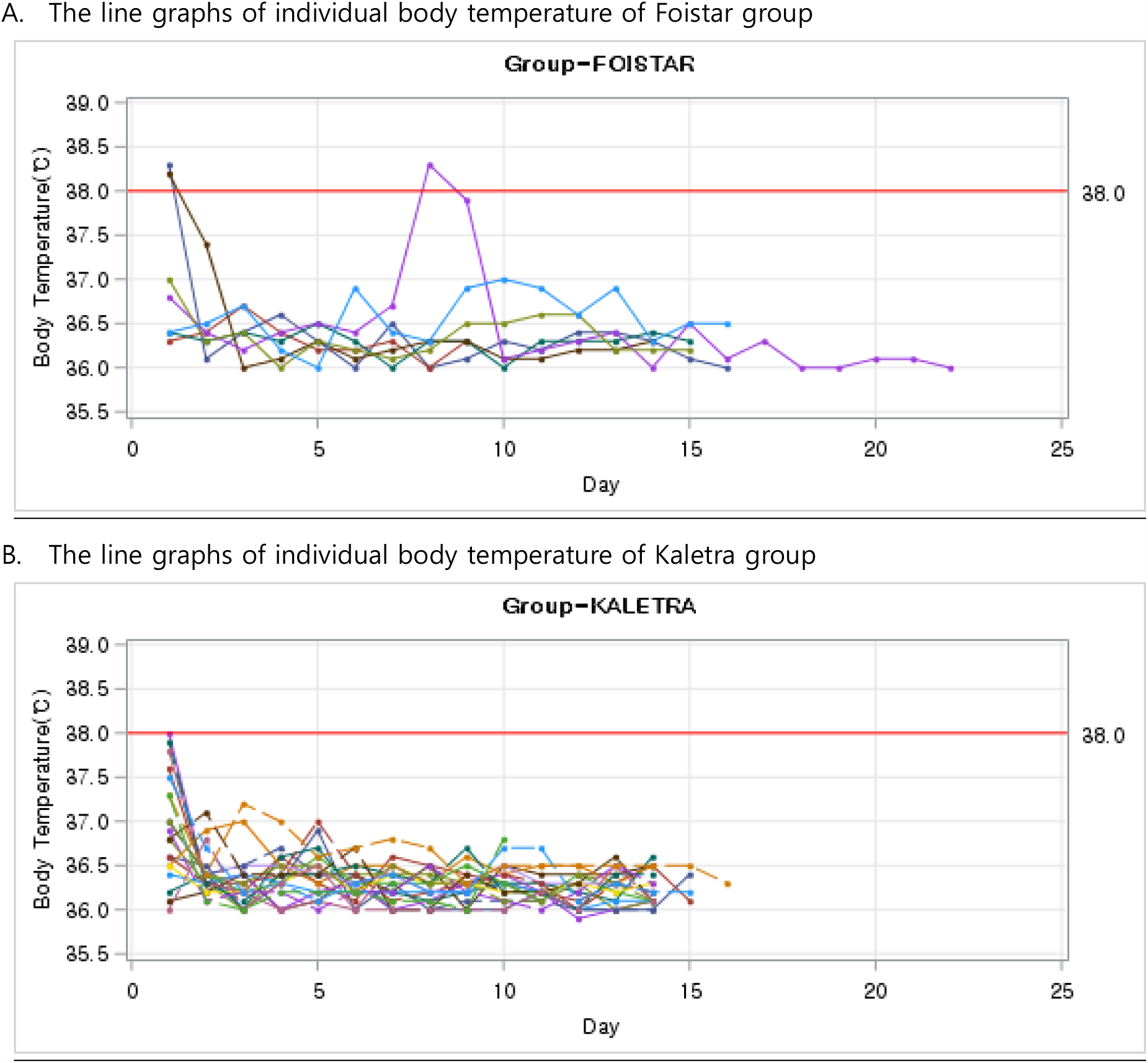
Body temperature for each patient during the hospitalization period.

The patient who became pyretic in Foistar group was administered with camostat one day before the event of fever and required high concentration of oxygen support. The fever was relieved on the following day.

### 3) Laboratory tests (CRP level, WBC count)

#### ➀ CRP

All seven patients in Foistar group had the elevated CRP levels (>0.4mg/dL) upon admission.

Among Kaletra group, two patients had the normal CRP levels upon the admission, and 18 patients had the elevated CRP levels, while two patients did not proceed the laboratory tests at the admission date. The trends of CRP levels during the hospitalization period in all patients were as follows. (Figure 2.)

**Figure 2.**
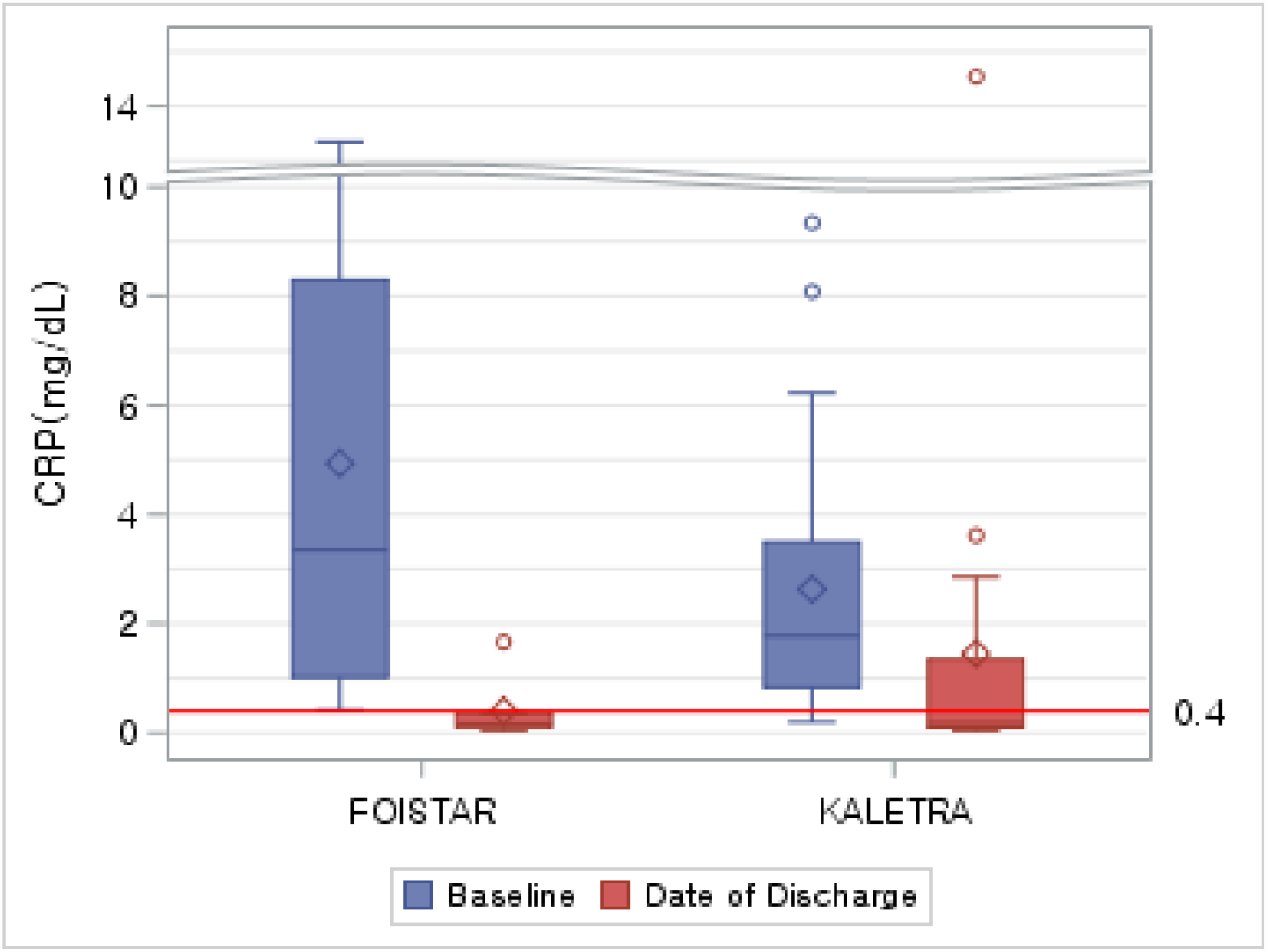
Changes of CRP levels between Foistar group and Kaletra group.

**Figure 3.**
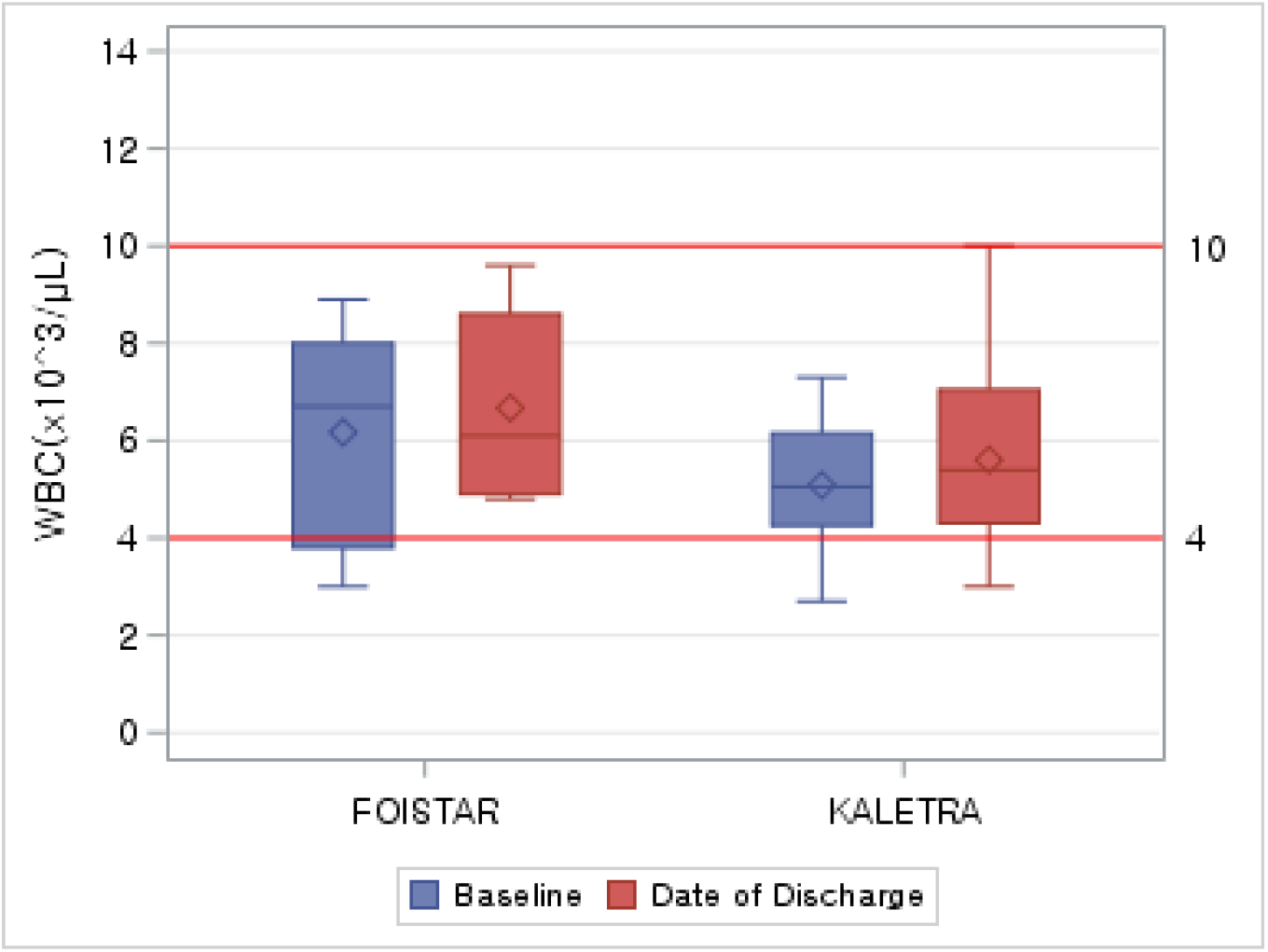
WBC for each group at date of discharge.

We were able to observe a probable association of decrease in CRP level with Foistar use. Six patients (85.71%) in Foistar group have achieved the normal CRP level, while 11 patients (50.00%) have achieved the normal CRP level in Kaletra group. Only was one of the patients (5.00%) in Kaletra group who had the normal CRP level upon the admission able to maintain the level under 0.4mg/dL. (Table 2.)

**Table 2.** Proportion of Improvement(≤0.4mg/dL) in CRP at date of discharge.

|  | Foistar Group<br>(N=7) | Kaletra Group<br>(N=20) | Total<br>(N=27) |
| --- | --- | --- | --- |
| **Proportion of Improvement($\leq 0.4$mg/dL) in CRP at date of discharge, n(%)** |  |  |  |
| $\leq 0.4$mg/dL at baseline → $\leq 0.4$mg/dL at Date of Discharge | 0 (0.00) | 1 (5.00) | 1 (3.70) |
| $\leq 0.4$mg/dL at baseline → $> 0.4$mg/dL at Date of Discharge | 0 (0.00) | 1 (5.00) | 1 (3.70) |
| $> 0.4$mg/dL at baseline → $\leq 0.4$mg/dL at Date of Discharge | 6 (85.71) | 11 (55.00) | 17 (62.96) |
| $> 0.4$mg/dL at baseline → $> 0.4$mg/dL at Date of Discharge | 1 (14.29) | 7 (35.00) | 8 (29.63) |

\* CRP Normal Range: $\leq 4.0$ mg/dL
\* No CRP result at baseline (Control group: 2 subjects).
\* If there is no value at the date of discharge, the last value before discharge was used.

#### ➁ WBC

Among Foistar group, 5 patients had the normal WBC count (4 -10 x 10^3^/uL), and 2 patients had the abnormal WBC count upon the admission. Among Kaletra group, 17 patients had the normal WBC counts while 3 patients had the abnormal WBC counts and the rest of 2 patients did not proceed the laboratory tests upon the admission.

The number of patients who had the abnormal WBC counts upon the admission became normal was 2 in Foistar group, and 1 in Kaletra group. The number of patients who had normal WBC counts upon the admission maintained normal was 5 in Foistar group, and 15 in Kaletra group.

#### ➂ Potassium

All 7 patients in Foistar group had the normal serum potassium level (3.5-5.5 mg/dL) upon the admission. Among Kaletra group, 19 patients had the normal serum potassium level, one of them was abnormal. The rest of 2 patients did not proceed the laboratory tests, therefore, they were excluded from the analysis

The all seven patients (100%) who had the normal potassium level in Foistar group were able to maintain their normal potassium level until discharge while 18 patients (90.00%) in the control group maintained their normal potassium levels. One of the patients (5.00%) who had the normal potassium level upon the admission in Kaletra group has become abnormal. Among the patients who had the abnormal potassium level in the control group, 1 patient (5.00%) has remained abnormal until discharge.

### 4) Event of diarrhea

An event of diarrhea during the hospitalization period was chart-reviewed. No patient in Foistar group had an event of diarrhea, while 9 patients (40.91%) had one or more events of diarrhea in Kaletra group until the discharge.

## Conclusion

Among Foistar group, the event of fever was observed in 1 patient, and none was observed in Kaletra group during the hospitalization period.

We have observed a possible advantage of Foistar on the anti-inflammatory effect against COVID-19, given 6 patients out of 7 patients (85.71%) from Foistar group who exhibited elevated CRP levels (CRP >0.4mg/dL) upon the admission have controlled their CRP levels to the normal range. Meanwhile, we concluded that the comparison on fever control, and WBC counts was considered inappropriate due to the significant difference in the baseline between two groups.

In addition, we could suggest further safety upon Foistar use as none of the patients have displayed hyperkalemia. a known adverse reaction of Foistar.^25^ On the other hand, we observed that the use of Kaletra was likely associated with an event of diarrhea. More than one event of diarrhea was observed from 9 patients out of 22 patients (40.91%) from Kaletra group, while none was found in Foistar group.

In conclusion, this study was able to demonstrate a probable association in fever control and the anti-inflammatory effects with the administration of Foistar in COVID-19 patients^26,27^. Further, the feasible safety profile of Foistar was observed in this study. Thus, the clinical use of Foistar is expected to have a favorable association with managing symptoms and clinical manifestations of COVID-19 such as fever, inflammation, etc. Although a further large-scale clinical trial with diverse clinical endpoints and the evaluation of virus titer is warranted, our study outcomes may positively encourage the use of Foistar as a treatment option for COVID-19.

## Discussion

Kaletra (lopinavir/ritonavir) is currently being used in COVID-19 patients as a possible therapeutic drug. However, it has been known to cause adverse reactions in the gastrointestinal tract such as diarrhea, nausea, and vomiting. In fact, the patients from the control group in this study have complained of diarrhea as well with minimal effectiveness against COVID-19 infection.

On the other hand, Foistar, which has recently been considered COVID-19 treatment, has established a stable safety profile since it has been marketed for a long period of time. For instance, hyperkalemia, which is one of the most common adverse reactions^25^, has not even occurred in this study. In addition, this is the first ever study where the anti-inflammatory effects of Foistar were confirmed in COVID-19 patients, thus, the results led to conclude that Foistar can be beneficial in the clinical settings for the mild COVID-19 patients. A future confirmatory large-scale prospective clinical trial of Foistar in COVID-19 infection is warranted to further ascertain its efficacy as COVID-19 treatment.

## Data Availability

All data referred in the manuscript are provided in the results and supplementary materials.

## Notes

### Competing Interest Statement

The authors have declared no competing interest.

### Clinical Trial

The study is not required to be registered in an internationally recognized trial since this is a retrospective cohort study.

### Funding Statement

All authors have nothing to disclose with respect to conflicts of interest or funding.

### Author Declarations

The present study protocol was reviewed and approved by the Institutional Review Board of Seoul Medical Center (approval No. 2020-11-039). The a signed consent was waived by IRB, since this retrospective study was considered a minimal risk study to the patients.

