## Supplementary Materials for "Foistar®(Camostat mesylate) associated with the significant decrease in CRP levels compared to Kaletra®(Lopinavir/Ritonavir) treatment in Korean mild COVID-19 pneumonic patients"

**1. Laboratory Test Results**

**1) CRP**

Figure 4. CRP levels for each patient during hospitalization

A. The individual CRP levels of Foistar group

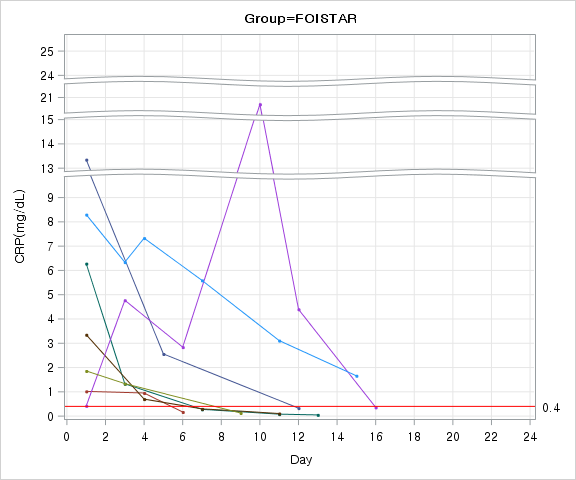

A. The individual CRP levels of Kaletra group

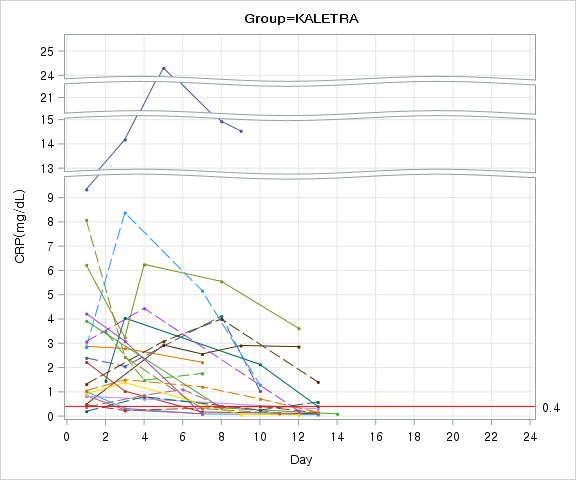

**2) WBC**

Figure 5. WBC for each patient during hospitalization

A. The individual WBC counts of Foistar group

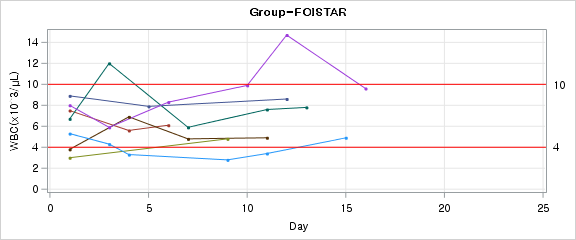

B. The individual WBC counts of Kaletra group.

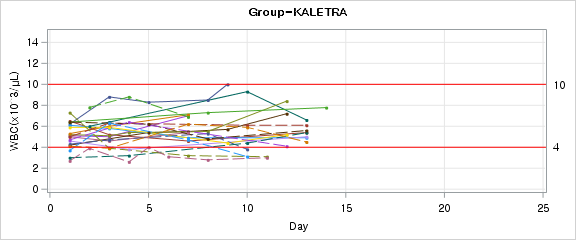

Six patients (85.71%) in the test group have achieved the normal CRP level, while 11 patients (50.00%) have achieved the normal CRP level in the control group. Only was one of the patients (5.00%) in the control group who had the normal CRP level upon the admission able to maintain the level under 0.4mg/dL. (Table 3.)

Table 3. Proportion of Improvement (Normal: 4-10) in WBC at date of discharge

|  | **Test Group** | | **Control Group** | **Total** |
| --- | --- | --- | --- | --- |
|  | **(N=7)** | | **(N=20)** | **(N=27)** |
| **Proportion of Improvement(Normal: 4-10) in WBC at date of discharge, n(%)** | | | | |
| Normal at baseline→ Normal at Date of Discharge | | 5 (71.43) | 15 (75.00) | 20 (74.07) |
| Normal at baseline→> Abnormal at Date of Discharge | | 0 (0.00) | 2 (10.00) | 2 (7.41) |
| Abnormal at baseline→ Normal at Date of Discharge | | 2 (28.57) | 1 (5.00) | 3 (11.11) |
| Abnormal at baseline→ Abnormal at Date of Discharge` | | 0 (0.00) | 2 (10.00) | 2 (7.41) |

* WBC Normal Range: 4-10

* No WBC result at baseline (Control group: 2 subjects).

* If there is no value at the date of discharge, the last value before discharge was used.

**2. WHO Ordinal Scale for Clinical Improvement**^24^

| *Ordinal Scale for Clinical Improvement* | | |
| --- | --- | --- |
| **Patient Status** | **Descriptor** | **Score** |
| **Uninfected** | No clinical or virological evidence of infection | 0 |
| **Ambulatory** | No limitation of activities | 1 |
|  | Limitation of activities | 2 |
| **Hospitalized Mild Disease** | Hospitalized, no oxygen therapy | 3 |
|  | Oxygen by mask or nasal prongs | 4 |
| **Hospitalized Severe Disease** | Non-invasive ventilation or high-flow oxygen | 5 |
|  | Intubation and mechanical ventilation | 6 |
|  | Ventilation+additional organ support-ECMO, CRRT, pressors | 7 |
| **Dead** | Death | 8 |

*COVID-19 Therapeutic Trial Synopsis(WHO R&D Blueprint, 2020.2.18.)*
